# Clinical and economic evaluation of a proteomic biomarker preterm birth risk predictor: Cost-effectiveness modeling of prenatal interventions applied to predicted higher-risk pregnancies within a large and diverse cohort

**DOI:** 10.1101/2021.09.08.21262940

**Authors:** Julja Burchard, Glenn R. Markenson, George R. Saade, Louise C. Laurent, Kent D. Heyborne, Dean V. Coonrod, Corina N. Schoen, Jason K. Baxter, David M. Haas, Sherri A. Longo, Scott A. Sullivan, Sarahn M. Wheeler, Leonardo M. Pereira, Kim A. Boggess, Angela F. Hawk, Amy H. Crockett, Ryan Treacy, Angela C. Fox, Ashoka D. Polpitiya, Tracey C. Fleischer, Thomas J. Garite, J. Jay Boniface, John A. F. Zupancic, Gregory C. Critchfield, Paul E. Kearney

## Abstract

**Objectives:** Preterm birth occurs in more than 10% of U.S. births and is the leading cause of U.S. neonatal deaths, with estimated annual costs exceeding $25 billion USD. Using real-world data, we modeled the potential clinical and economic utility of a prematurity-reduction program comprising screening in a racially and ethnically diverse population with a validated proteomic biomarker risk predictor, followed by case management with or without pharmacological treatment.

**Methods:** The ACCORDANT microsimulation model used individual patient data from a prespecified, randomly selected sub-cohort (N=847) of a multicenter, observational study of U.S. subjects receiving standard obstetric care with masked risk predictor assessment (TREETOP; NCT02787213). All subjects were included in three arms across 500 simulated trials: standard of care (SoC, control); risk predictor/case management comprising increased outreach, education and specialist care (RP-CM, active); and risk predictor/case management with pharmacological treatment (RP-MM, active). In the active arms, only subjects stratified as higher-risk by the predictor were modeled as receiving the intervention, whereas lower-risk subjects received standard care. Higher-risk subjects’ gestational ages at birth were shifted based on published efficacies, and dependent outcomes, calibrated using national datasets, were changed accordingly. Subjects otherwise retained their original TREETOP outcomes. Arms were compared using survival analysis for neonatal and maternal hospital length of stay, bootstrap intervals for neonatal cost, and Fisher’s exact test for neonatal morbidity/mortality (significance, *p*<0.05).

**Results:** The model predicted improvements for all outcomes. RP-CM decreased neonatal and maternal hospital stay by 19% (*p*=0.029) and 8.5% (*p*=0.001), respectively; neonatal costs’ point estimate by 16% (*p*=0.098); and moderate-to-severe neonatal morbidity/mortality by 29% (*p*=0.025). RP-MM strengthened observed reductions and significance. Point estimates of benefit did not differ by race/ethnicity.

**Conclusions:** Modeled evaluation of a biomarker-based test-and-treat strategy in a diverse population predicts clinically and economically meaningful improvements in neonatal and maternal outcomes.

**Plain language summary:** Preterm birth, defined as delivery before 37 weeks’ gestation, is the leading cause of illness and death in newborns. In the United States, more than 10% of infants is born prematurely, and this rate is substantially higher in lower-income, inner-city and Black populations. Prematurity associates with substantially increased risk of short- and long-term medical complications and can generate significant costs throughout the lives of affected children. Annual U.S. health care costs to manage short- and long-term prematurity complications are estimated to exceed $25 billion.

Clinical interventions, including case management (increased patient outreach, education and specialist care), pharmacological treatment and their combination, can provide benefit to pregnancies at higher risk for preterm birth. Early and sensitive risk detection, however, remains a challenge.

We have developed and validated a proteomic biomarker risk predictor for early identification of pregnancies at increased risk of preterm birth. The ACCORDANT study modeled treatments with real-world patient data from a racially and ethnically diverse U.S. population to compare the benefits of risk predictor testing plus clinical intervention for higher-risk pregnancies versus no testing and standard care. Measured outcomes included neonatal and maternal length of hospital stay, associated costs and neonatal morbidity and mortality. The model projected improved outcomes and reduced costs across all subjects, including ethnic and racial populations, when predicted higher-risk pregnancies were treated using case management with or without pharmacological treatment. The biomarker risk predictor shows high potential to be a clinically important component of risk stratification for pregnant women, leading to tangible gains in reducing the impact of preterm birth.

## Introduction

Preterm birth (PTB), defined as delivery earlier than 37 weeks’ gestational age (GA), occurs in approximately 10% of all births and is the leading cause of neonatal deaths in the United States [1,2]. In addition to neonatal morbidity and mortality, the economic impact of PTB is enormous, exceeding $25 billion USD annually [3]. The direct medical care burden, accounting for most of these costs, increases markedly as GA at birth decreases.

Effective intervention to ameliorate PTB and its sequelae requires earlier identification of at-risk pregnancies. A history of prior spontaneous PTB (sPTB) is a traditional predictor of PTB but is seen in only approximately 4% of all U.S. pregnancies and predicts only 11% of all sPTBs [4,5]. Similarly, short cervical length is a widely used predictor of sPTB but occurs in only 2% of all U.S. pregnancies and accounts for only 6% of all sPTBs beyond those identified by prior history [6,7]. Furthermore, the use of prior sPTB as a risk factor is not applicable in first pregnancies, while measurement of cervical length requires additional health care resources. Beyond these clinical risk factors, it is well documented that socioeconomically challenged and ethnic and racial minority populations are at substantially greater risk, with many factors in these groups contributing to increased risk [3]. In particular, PTB rates in the United States in 2020 were 9.1% among non-Hispanic White individuals, 9.8% in Hispanic individuals, and 14.4% among non-Hispanic Black individuals [2]. There is a clear need for sensitive predictive approaches that apply to all at-risk individuals.

Biomarker-based risk stratification tools offer a precision diagnostic approach to predict increased risk of PTB, particularly among those who show no apparent clinical signs, well in advance of delivery [8]. In the prospective Proteomic Assessment of Preterm Risk study (PAPR; NCT01371019), Saade et al. [9] described and clinically validated a novel serum proteomic biomarker risk predictor for sPTB that combines the expression ratio of insulin-like growth factor-binding protein 4 (IBP4) to sex hormone-binding globulin (SHBG) with clinical variables. The mass spectrometry assay measuring the IBP4/SHBG ratio has been analytically validated [10]. This risk predictor, which identifies 75% of sPTBs, has been reported also to be predictive of indicated PTB [11], and in early clinical utility study the test was shown to benefit patients with all-cause PTB [12]. The PAPR analysis also established a predictive biomarker threshold score that significantly stratifies premature from later gestational ages at birth and corresponds to a 15% risk – double the average risk of sPTB across U.S. singleton pregnancies [3]. This threshold was validated in a secondary analysis [13] of the independent, prospective Multicenter Assessment of a Spontaneous Preterm Birth Risk Predictor study (TREETOP); NCT02787213) [14], in which the sensitivity of identifying preterm pregnancies was demonstrated to be 88%.

Identifying PTB risk with higher sensitivity than can be achieved using clinical risk factors alone enables the targeting of more at-risk patients with interventions under active study, including intensive case management (increased outreach, PTB education and specialist care) [15-17], pharmaceutical treatment [4,6] or their combination [12]. The purpose of the current analysis, named ACCORDANT (Analyses aCross Congruent studies ReDucing Adverse pregNancy ouTcomes), was to assess the cost effectiveness of two test-and-treat strategies relative to standard care from a healthcare payer perspective, modeling differences in outcomes including neonatal and maternal lengths of hospital stay (LOS), neonatal costs, and neonatal morbidity and mortality in a diverse U.S. pregnant population. A secondary analysis of the TREETOP study, ACCORDANT used proteomic biomarker risk predictor results to stratify subjects into PTB risk groups, then applied models of existing clinical interventions to individuals predicted to be at higher risk for premature delivery.

## Methods

ACCORDANT was conducted according to a prespecified plan, with the components listed below. A study overview appears in **Figure 1**. The analysis employed a microsimulation of pregnant individuals from a healthcare payer perspective, assessed from the second trimester of pregnancy through post-delivery maternal and neonatal discharge. The rationale for this methodology was to assess the range of effects on clinical and economic outcomes of modeled interventions on an actual population of pregnant women determined to be elevated risk by the risk predictor. This report was prepared in alignment with Consolidated Health Economic Evaluation Reporting Standards (CHEERS) 2022 guidance [18,19]. All costs were measured and reported in 2019 U.S. dollars.

**Figure 1.**
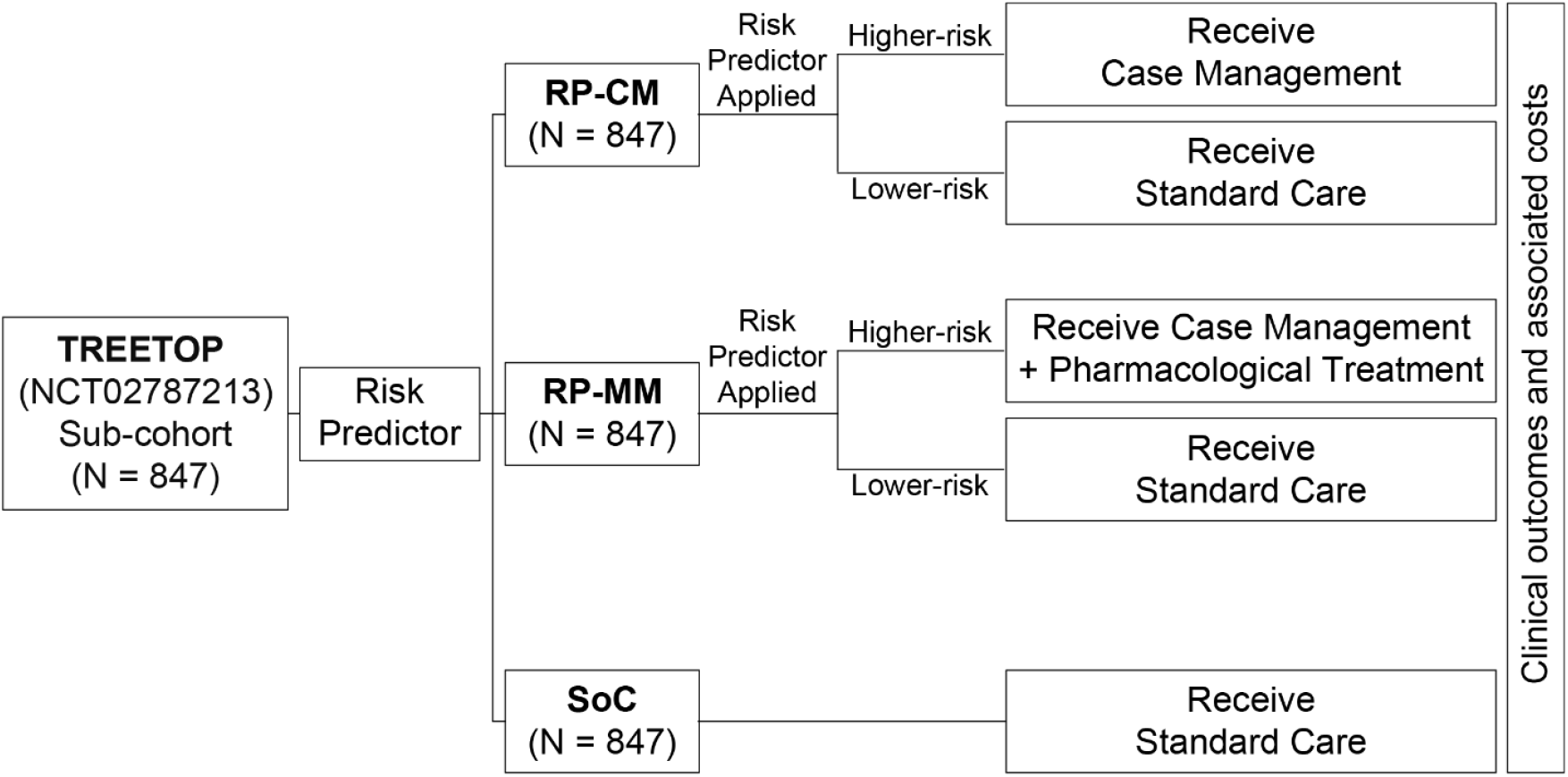
Schematic overview of the ACCORDANT study. ACCORDANT subjects (N = 847) were routed to three study arms: standard care (SoC) control; risk predictor plus case management intervention (RP-CM); or risk predictor plus case management intervention with pharmacological treatment (RP-MM). Those in the RP-CM and RP-MM arms were stratified into higher-risk and lower-risk groups based on their proteomic biomarker risk predictor scores, using a previously validated threshold. Subjects predicted to be at higher PTB risk were modeled to receive case management alone (RP-CM) or case management with pharmacological treatment (RP-MM). Lower-risk subjects in the RP-CM and RP-MM arms received standard care. The gestational ages at birth of higher-risk subjects were shifted according to published intervention efficacies. Associated outcomes were calculated for the treated groups and for subjects with missing or truncated data in both arms. Outcomes were compared between active arms and the control arm, as a whole and between the most severely affected 10% of each arm by outcome.

### ACCORDANT study population

The study population comprised a sub-cohort (N = 847) of subjects enrolled in the Multicenter Assessment of a Spontaneous Preterm Birth Risk Predictor study (TREETOP; NCT02787213) [14]. TREETOP was an observational study of pregnant women across the United States who were deemed not to be at higher risk for PTB by traditional clinical methods. Study subjects were independent from those used to develop the risk predictor [9] and assess intervention efficacy [12,15-17], and they had not been used in previous modeling studies. Subjects represented a geographically, ethnically, and racially diverse cohort of women receiving care at 18 U.S. tertiary care centers. Race and ethnicity were self-identified by participants, and diversity in the cohort was ensured by intentional over-enrollment of Black (19.3% versus 16.9% nationally) and Hispanic (39.2% versus 19.9% nationally) individuals [20]. Clinical management of patients was via standard of care, as risk predictor test results were masked. Outcomes were observed in TREETOP through the end of the neonatal period (28 days). Subjects were selected randomly by a third-party statistician as part of a planned first-phase analysis of 34% of TREETOP study completers and reflect the demographic makeup of the overall study. The TREETOP study was approved by the Institutional Review Board at each participating study center, and all participants provided written informed consent prior to enrollment in the study.

### Study arms

Three arms were modeled: 1) a control arm of standard care (SoC) as practiced in TREETOP; 2) an active arm of risk predictor stratification of pregnancies to case management (RP-CM) designated as the base case; and 3) an active arm of risk predictor stratification of pregnancies to multimodal treatment including case management and pharmacological intervention (RP-MM). Both active arms reflected current clinical practice scenarios and modeled the effects of care as a shift in GA at birth. Outcomes were assigned dependent upon GA at birth, actual or modeled depending on the study arm and risk predictor test result.

Studies of interventions to ameliorate PTB risk and effects report changes in distribution of GA at birth with treatment [12,15-17]. These distributional changes can be accurately quantified as shifts in GA at birth upon treatment that depend on the untreated GA, with larger GA shifts amongst the earliest births and decreasing shifts as GA increases. Therefore, the analysis did not anticipate a significant change in overall PTB rate, nor a shift in mean or median GA at birth across the study population, as most births were term births and were unaffected. The RP-CM and RP-MM arm models, derived from such studies, respectively incorporated expected literature-based probabilistic shifts in GA at birth calculated as a function of the GA at birth observed without intervention in TREETOP. Specifically, the RP-CM and RP-MM models provided GA-specific means and standard deviations of shifts in GA at birth with treatment, relative to TREETOP GA at birth. Specifically, for each treated subject, the observed GA at birth was shifted by an amount selected randomly from a normal distribution. The mean and standard deviation of the distribution were set by sigmoid functions (**Supplemental Figure 1**) fitted to tables of numbers of deliveries by GA for untreated and treated women in the intervention studies. The normal distribution derived for each subject represented the expected range of changes in GA at birth with RP-CM or RP-MM strategies compared with SoC, representing the uncertainty in assigning values to the variable shift in GA. Coefficients used in the sigmoid functions are specified in **Table 1**. For illustration, magnitudes of mean shift in GA at birth in each active arm are presented for weeks 28 to 36 in **Figure 2** and **Supplemental Table 2**.

**Table 1.** Summary of ACCORDANT assumptions.

| **Input** | **Type** | **Center** | **Spread** | **Distribution** | **Reference** |
| --- | --- | --- | --- | --- | --- |
| **Size of population** | Demographic | As observed | As observed | N/A | Markenson [14] |
| **Composition of population** | Demographic | As observed | As observed | N/A | Markenson [14] |
| **Test performance** | Clinical | As observed | As observed | N/A | Markenson [14] |
| **Untreated GA at birth** | Clinical | As observed | As observed | N/A | Markenson [14] |
| **Untreated neonatal LOS** | Clinical | As observed | As observed | N/A | Markenson [14] |
| **Untreated maternal LOS** | Clinical | As observed | As observed | N/A | Markenson [14] |
| **Untreated neonatal morbidity and mortality** | Clinical | As observed | As observed | N/A | Markenson [14] |
| **Multimodal intervention** | Clinical | $max\_m = 9.5$ weeks<br>$mid\_m = 30.75$ weeks<br>$rate\_m = -1/2.3^\dagger$ | $max\_SD = 3.09$ weeks<br>$mid\_SD = 30.13$ weeks<br>$rate\_SD = -1/2.9^\dagger$ | Three-parameter sigmoid | Branch [12] |
| **Case management intervention** | Clinical | $max\_m = 5.25$ weeks<br>$mid\_m = 30.75$ weeks<br>$rate\_m = -1/2.1^\dagger$ | $max\_SD = 3.09$ weeks<br>$mid\_SD = 30.13$ weeks<br>$rate\_SD = -1/2.9^\dagger$ | Three-parameter sigmoid | Stankaitis [15]<br>Newman [16]<br>Newnham [17] |
| **Treated neonatal LOS for GA<35 weeks (MIX)** | Clinical | GA-dependent as published;<br>80% of mean applied to gamma, 20% to lognormal deviates | GA-dependent as published;<br>60% of variance applied to gamma, 40% to lognormal deviates | Method of moments:<br>replicating parametric and non-parametric summary statistics | MIX [21] |
| **Treated neonatal LOS for GA $\geq 35$ weeks (MIX)** | Clinical | GA-dependent as published;<br>10% of mean applied to gamma, 90% to lognormal deviates | GA-dependent as published;<br>90% of variance applied to gamma, 10% to lognormal deviates | Method of moments:<br>replicating parametric and non-parametric summary statistics | MIX [21] |
| **Treated neonatal LOS for GA<35 weeks (COM)** | Clinical | GA-dependent as published;<br>85% of mean applied to gamma, 15% to lognormal deviates | GA-dependent as published;<br>60% of variance applied to gamma, 40% to lognormal deviates | Method of moments:<br>replicating parametric and non-parametric summary statistics | COM [22] |
| **Treated neonatal LOS for GA $\geq 35$ weeks (COM)** | Clinical | GA-dependent as published;<br>15% of mean applied to gamma, 85% to lognormal deviates | GA-dependent as published;<br>90% of variance applied to gamma, 10% to lognormal deviates | Method of moments:<br>replicating parametric and non-parametric summary statistics | COM [22] |
| **Treated maternal LOS for GA <35 weeks** | Clinical | GA-dependent as published; 20% of mean applied to gamma, 80% to lognormal deviates | GA-dependent as published; 60% of variance applied to gamma, 40% to lognormal deviates | Method of moments: replicating parametric and non-parametric summary statistics | MIX [21] |
| **Treated maternal LOS for GA ≥35 weeks** | Clinical | GA-dependent as published; 20% of mean applied to gamma, 80% to lognormal deviates | GA-dependent as published; 80% of variance applied to gamma, 20% to lognormal deviates | Method of moments: replicating parametric and non-parametric summary statistics | MIX [21] |
| **Treated neonatal morbidity and mortality** | Clinical | See Supplemental Table 1 |  | Multinomial | Markenson [14] |
| **Treated or untreated neonatal LOS for cases of perinatal mortality** | Clinical | One day longer than maximum observed | 0 | N/A | Branch [12] |
| **Neonate cost for cases of perinatal mortality** | Economic | \$89,069 | \$0 | Point estimate: average observed | MIX [21] |
| **Neonate cost** | Economic | GA-dependent cost per day as published applied to neonatal LOS | As for neonatal LOS | Method of moments: blend of gamma and lognormal | MIX [21] |
| **Neonate cost** | Economic | GA-dependent cost per day as published applied to neonatal LOS | As for neonatal LOS | Method of moments: blend of gamma and lognormal | COM [22] |
| **Neonate cost adjustment** | Economic | MIX [21] (12/2017) to TREETOP (4/2019): 1.026; COM [22] (2016) to TREETOP (4/2019): 1.049 | n/a | N/A | FRED [28] |
| **Progesterone cost** | Economic | \$20 | \$0 | Point estimate | Fried [24]<br>Cohen [25]<br>Cahill [26]<br>Kowalski [27] |
| **Case management cost** | Economic | \$53 | \$0 | Point estimate | Estimated from CMS physician fee schedule for PREVENT [12] protocol care |

<sup>†</sup>*max<sub>m</sub>* and *max<sub>SD</sub>* are the means and standard deviations for the upper asymptotes; *mid<sub>m</sub>* and *mid<sub>SD</sub>* are the means and standard deviations for the inflection points; and *rate<sub>m</sub>* and *rate<sub>SD</sub>* are the means and standard deviations for the slopes of the three
parameter sigmoid functions.
Abbreviations. CMS, Centers for Medicare & Medicaid Services; COM, U.S. commercial payer dataset; GA, gestational age; LOS, hospital length of stay; MIX, U.S. state-based dataset with a mix of insurance payers.

**Table 2.**
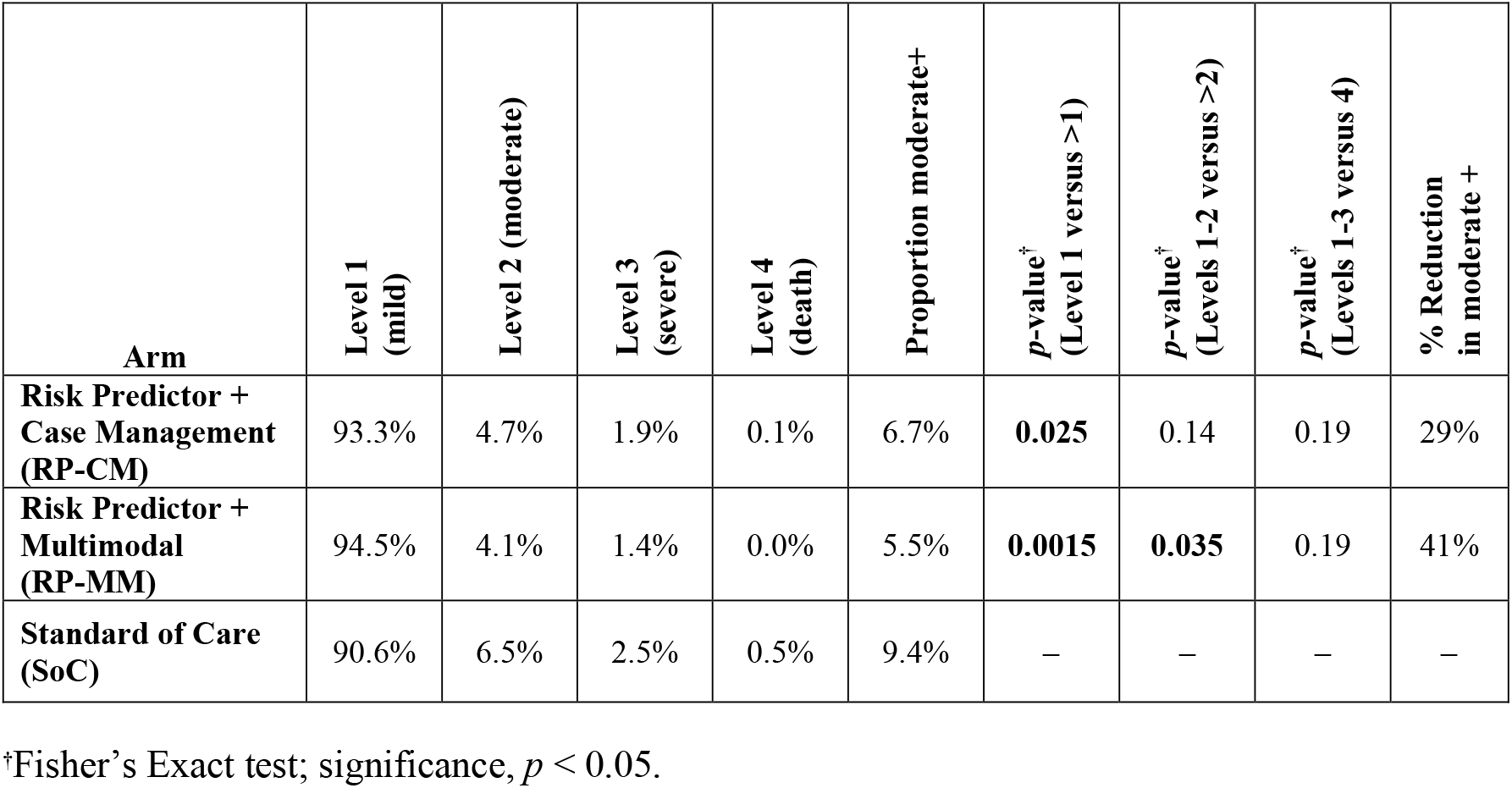
Impact of test-and-treat strategies on neonatal morbidity and mortality. Comparisons were made between the control arm and the two active arms of the proportion of subjects at or below versus above each elevated level of the neonatal morbidity and mortality index. The percent reduction in levels 2 or higher is shown for each active arm. Significant *p*-values are indicated in boldface type.

| Arm | Level 1<br>(mild) | Level 2 (moderate) | Level 3<br>(severe) | Level 4<br>(death) | Proportion moderate+ | *p*-value<sup>†</sup><br>(Level 1 versus >1) | *p*-value<sup>†</sup><br>(Levels 1-2 versus >2) | *p*-value<sup>†</sup><br>(Levels 1-3 versus 4) | % Reduction<br>in moderate + |
| --- | --- | --- | --- | --- | --- | --- | --- | --- | --- |
| **Risk Predictor +**<br>**Case Management**<br>**(RP-CM)** | 93.3% | 4.7% | 1.9% | 0.1% | 6.7% | **0.025** | 0.14 | 0.19 | 29% |
| **Risk Predictor +**<br>**Multimodal**<br>**(RP-MM)** | 94.5% | 4.1% | 1.4% | 0.0% | 5.5% | **0.0015** | **0.035** | 0.19 | 41% |
| **Standard of Care**<br>**(SoC)** | 90.6% | 6.5% | 2.5% | 0.5% | 9.4% | — | — | — | — |

<sup>†</sup>Fisher's Exact test; significance, $p < 0.05$ .

**Figure 2.**
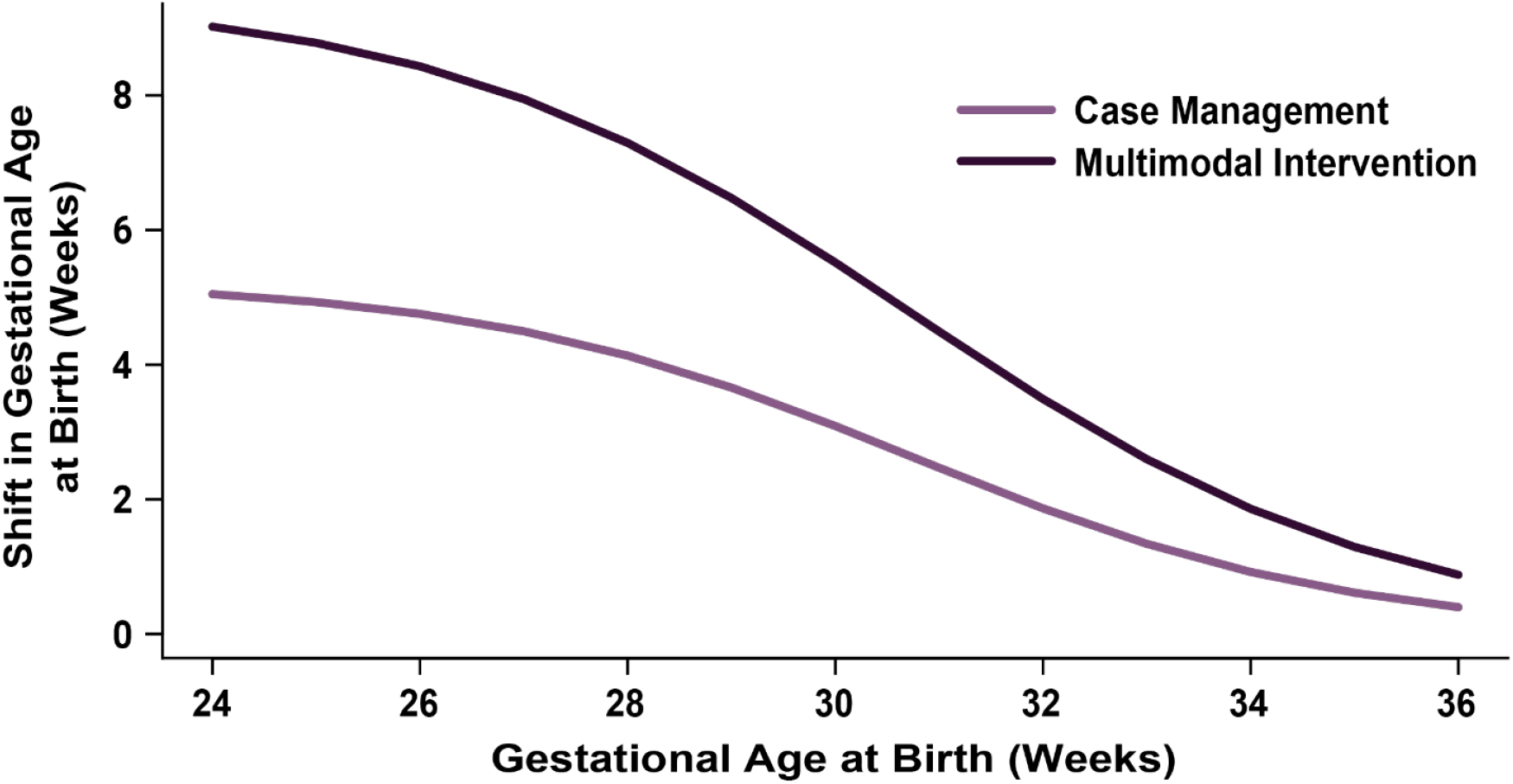
Shifts in gestational ages at birth applied to higher-risk subjects modeled to receive case management (increased outreach, preterm education and specialist care) or multimodal intervention (case management with pharmaceutical treatment). Shifts were based on published intervention efficacies.

### Outcomes

Measured outcomes included maternal and total neonatal hospital length of stay (LOS), (comprising both neonatal intensive care unit and nursery care), neonatal costs, and neonatal morbidity and mortality. Data sources used for simulated outcomes are detailed as follows.

### Model inputs

Input data on GA at birth and outcomes in the SoC arm were sourced from TREETOP. Clinical data were recorded electronically, monitored onsite, and were subject to predefined source document verification. PTB outcomes were adjudicated by an independent panel of three physicians.

TREETOP was the source of a multinomial model of neonatal morbidity and mortality dependent on GA at birth. California and national data (defined below as MIX and COM, respectively) provided the dependence of maternal and neonatal hospital LOS and neonatal cost on GA at birth [21,22]. Efficacy of interventions for PTB reduction was derived from published studies of stratification of higher-risk pregnancies to case management (RP-CM) [12,15-17] or to multimodal treatment combining case management with pharmacologic interventions (RP-MM) [12].

Each microsimulation subject had been enrolled in TREETOP with records of maternal and neonatal outcomes adjudicated based on prevailing definitions. Each subject was assigned her own observed TREETOP outcomes unless she was stratified to receive interventions in the RP-CM or RP-MM arms. In instances when a datapoint was missing for any subject, a simulated outcome was assigned.

### Risk stratification

In TREETOP, the risk predictor was assayed in all subject samples as they were collected during GA weeks 19^1/7^ – 20^6/7^, using a validated and standardized laboratory process [9,10] that was consistent with clinical intended use. In ACCORDANT, each subject’s laboratory measurement was compared to the validated risk threshold at about twice the baseline rate in U.S. singleton pregnancies, or 15% risk [9,13], stratifying subjects to higher and lower-risk groups.

### Simulated interventions

In the RP-CM and RP-MM arms, interventions were applied to subjects whose risk predictor scores indicated higher PTB risk. RP-CM and RP-MM subjects stratified as lower risk received standard care. The RP-CM intervention model was based on results from three controlled studies [15-17] on the effect of case management applied to pregnancies stratified to treatment, with higher-risk status determined by pre-specified clinical and/or demographic risk factors. Shifts in GA at birth for the RP-CM model, as indexed by untreated GA at birth, were derived using the metafor R package [23]. The RP-MM intervention model was based on the observed effect on GA at birth of applying case management with pharmaceutical treatment (17-α-hydroxyprogesterone caproate). The intervention effect model was fitted to data from Prediction and Prevention of Preterm Birth (PREVENT-PTB, NCT03530332), a randomized controlled trial [12] that followed women at otherwise low risk of PTB assigned to standard care or to risk predictor screening followed by multimodal interventions. Costs of case management were estimated from the CMS physician fee schedule for PREVENT-PTB protocol care **(Table 1)**. Costs of pharmaceutical treatment were as published [24-27].

### Neonatal and maternal length of hospital stay

After intervention and to replace missing or truncated TREETOP data, lengths of stay (LOS) were sourced from reports of LOS and associated costs stratified by GA two studies of large datasets collected in the U.S., one (MIX) based on California state data with a mix of payers [21] and one (COM) based on commercial payer data [22]. MIX and COM support generalizability as their datasets included over 1,000,000 and 750,000 pregnancies, respectively. Neonatal and maternal LOS were indexed by weeks of GA at birth and were mapped to microsimulation subjects based on GA.

### Neonatal costs

Neonatal costs associated with the birth hospital admission were generated from MIX [21] and COM [22] along with LOS, enabling reproduction of the published relationships between cost and LOS. The two datasets reported costs in U.S. dollars. Costs were adjusted to April 2019 values, corresponding to the end of the TREETOP study, using the Producer Price Index by Industry [28] then indexed by weeks of GA at birth and assigned to microsimulation subjects on this basis. The analysis modeled gross savings independent of test price.

### Neonatal morbidity and mortality

Assessment of neonatal morbidity and mortality was based on an established scoring system [6]. Affected infants were assigned a morbidity and mortality index score that increased from 1 (mild) to 2 (moderate) to 3 (severe) for each additional diagnosis of one of respiratory distress syndrome, bronchopulmonary dysplasia, intraventricular hemorrhage grade III or IV, all stages of necrotizing enterocolitis, periventricular leukomalacia or proven severe sepsis; with a score of 4 assigned to perinatal mortality. The scale used hospital stays to determine index scores if the LOS gave a higher score than did concomitant diagnoses: 1-4 days gave a score of 1, 5-20 days a score of 2, and >20 days a score of 3. For example, level 1 included neonates with at most one adverse outcome and up to four days of hospital stay. While the scale accounts for neonatal intensive care unit stays, total length of neonatal hospital stay was used here instead to leverage the universality of hospital admission and for greater simplicity in calculating hospital LOS versus level of care.

Microsimulation subjects received initial neonatal morbidity and mortality index scores based on TREETOP observations. Changes in index scores with intervention were based on multinomial distributions derived from TREETOP and indexed by GA at birth (**Supplemental Table 1**).

### Outcome measurement

For microsimulation subjects, measured outcomes were either within error ranges for the values observed in TREETOP if unaffected by intervention, mortality or absent data; otherwise, they were assigned as a function of GA at birth. Microsimulation of study arms generated GAs at birth for either the base distribution (SOC) or shifted distributions (RP-CM and RP-MM). Outcomes were derived from calibrated calculations as probabilistic functions of GA at birth.

Post-intervention LOS and missing or truncated TREETOP neonatal LOS information were simulated. TREETOP truncated collection of LOS for each mother and neonate at 28 days post-delivery, affecting measurements of neonatal but not maternal LOS available for ACCORDANT subjects.

Neonatal LOS was generated by drawing random samples from distributions developed from the COM and MIX data. For each GA at birth, the method of moments was used to derive blended gamma and lognormal distributions from published means and variances. Calibration to source data was confirmed by agreement with published nonparametric estimates. The distributions also were confirmed to be calibrated to the distributions of LOS in TREETOP up to the limit of data collection, reducing artifactual divergence of observed and simulated values.

Based on the published observation in COM [22] that late-preterm LOS distributions were shaped differently than were early-preterm distributions, distinct distributions were applied to early and late preterm GAs at birth **(Table 1)**. A random draw from either the MIX or COM distribution replaced LOS for subjects whose GA at birth was shifted by intervention and those with missing/truncated data. For subjects with no GA shift and with valid data, LOS was adjusted based on random draws relative to TREETOP observations by up to the estimated error of LOS measurement of ±1 day). Maternal LOS was determined similarly, except only MIX was used (maternal LOS was not available in COM).

Per-day neonatal costs were calculated using the GA-specific total cost and associated LOS in the birth admission separately for the MIX and COM. These per-diem costs were then applied to the LOS assigned to microsimulation subjects. Derivation of simulated cost from simulated LOS maintained the known tight association of cost and LOS and used the probabilistic distribution already generated by the neonatal LOS model. Subjects who underwent intervention also were assigned an intervention cost from weeks 22-35 for progesterone treatment and/or case management (see Table 1 for details).

ACCORDANT subjects were assigned an initial neonatal morbidity and mortality index score based on TREETOP observations. If the subject’s GA at birth changed, neonatal morbidity and mortality was updated based on multinomial distributions derived from TREETOP and indexed by GA at birth (**Supplemental Table 1**). As the neonatal morbidity and mortality index is an ordinal scale without numeric error, and as the interventions modeled have been shown to do no detectable harm [12,15-17], modeled neonatal morbidity and mortality was restricted to the original observation in untreated subjects and to values no greater than the original observation in treated subjects.

### Validation

Face validity was evaluated by expert opinion on model structure, data sources, time horizon and outcomes [29]. For internal validity, models were calibrated against their source data, code was checked by a separate programmer, and selection of statistical tests was subject to external review. Outlier values were traced and explored. In partially dependent external validation, cost results were compared to the decision-analytic model used in a separate risk predictor cost-effectiveness study [30], and results for RP-MM were compared to direct observation of PREVENT-PTB outcomes [12]. Sources for predictive validity are currently unavailable.

### Uncertainty and statistical analysis

In each simulated trial, all subjects participated in the RP-CM, RP-MM and SoC arms. The routing of all subjects to each arm allowed for demographically unbiased comparisons between arms. While the matched design enabled paired tests or tests of repeated measures, a conservative assumption of independence was applied across arms. For LOS and neonatal cost outcomes, MIX was chosen as the base case, while COM was employed in sensitivity analyses. To achieve robust results and assess variance, 500 trial simulations were performed. This number was chosen as a compromise between computational cost and stability of estimates. Effects were quantitated as the median and interquartile range (IQR) across the 500 trials. Analysis of demographic subgroups was underpowered and therefore was assessed as point estimates only.

Representations of uncertainty, standard deviations representing parameter uncertainty and assumptions for study implementation are detailed in **Table 1**. Intervention effects and intervention-dependent outcomes were generated as probabilistic distributions, capturing uncertainty of model and outcome parameters. In each active arm, uncertainty in GA shifts was modeled as random normal error. To address the variability of both long and short LOS, uncertainty was modeled as a mixture of lognormal and gamma distributions.

Significance of changes in neonatal and maternal LOS due to treatment was assessed by a prespecified time-to-event analysis. While multiple approaches give similar results, Cox proportional hazards regression *p*-values were reported here for the main effect of study arm, either in the study as a whole or in the top 10% of subjects per arm per outcome.

Significance of changes in the number of subjects in each neonatal morbidity and mortality level was assessed using a one-sided Fisher’s Exact test. A one-sided test was pre-specified, as the outcome of interest was reduction in occurrence of higher index scores of neonatal morbidity and mortality [31], and the modeled interventions have been shown to do no detectable harm [12,15-17].

Changes in cost with treatment were assessed by a prespecified bootstrap test of total costs, either in each arm as a whole or in the top 10% of neonates per arm by cost. As cost of prematurity is largely driven by the birth admission [31,32], uncertainty in LOS also provided uncertainty in cost. Cost distributions were generated from neonatal LOS distributions through multiplication by GA per-diem costs. Thus, cost uncertainty was derived directly from uncertainty in LOS.

Statistical analyses were performed in R (3.5.1 or higher; Microsoft R Application Network [33]). As this study was exploratory as per International Society for Pharmacoeconomics and Outcomes Research (ISPOR) classification [34], *p*-values < 0.05 were considered significant.

## Results

Of 847 subjects, 308 in the active arms (RP-MM: multimodal; RP-CM, case management) were identified as higher risk. Amongst the subjects predicted to be at higher risk for sPTB, the racial and ethnic distribution was 22.4% Black, 47.4% Hispanic, 24% White and 6.2% other groups; this represented a 1.5-fold increase in Black and 1.7-fold increase in Hispanic representation relative to White or other groups **(Supplemental Table 3)**. Application of interventions to higher-risk individuals resulted in median prolongation of gestation of less than one day (RP-CM, 0.3 days; RP-MM, 0.8 days) compared with controls (SoC arm). However, for the subgroup comprising the lowest 10% for GA at birth, the median prolongation of gestation was three days (IQR width, 0.64 days) for the RP-CM arm and eight days (IQR width, 0.34 days) for the RP-MM arm relative to the SoC arm.

**Figure 3** and **Supplemental Table 4** detail the impact of the two active arms on neonatal hospital LOS. Across all subjects, neonatal LOS was reduced in the RP-CM arm by 19% (MIX, 14-23%) and 22% (COM, 17-26%) and in the RP-MM arm by 26% (MIX, 22-29%) and 30% (COM, 25-33%) compared with the SoC arm. For subjects in the top 10% of neonatal LOS, stay was reduced in the RP-CM arm by 33% (MIX, 25-40%) and 34% (COM, 26-40%) and in the RP-MM arm by 46% (MIX, 41-52%) and 47% (COM, 41-52%) relative to the SoC arm. In all cases, regardless of the active arm or outcome dataset, neonatal LOS reductions versus the control arm were significant. Among self-identified Black individuals, median neonatal hospital LOS point estimates in the base-case scenario (RP-CM) were estimated to fall by 20% (MIX) and 25% (COM) across all subjects and by 35% (MIX) and 39% (COM) in the top 10% of neonatal LOS (**Figure 5**; **Supplemental Table 5**). Among self-identified Hispanic individuals, point estimates of neonatal LOS were seen to be reduced by 17% (MIX) and 21% (COM) across all subjects and by 31% (MIX) and 32% (COM) in the top 10% of neonatal LOS. Greater reductions in LOS were estimated when pharmacological treatment (RP-MM) was included.

**Figure 3.**
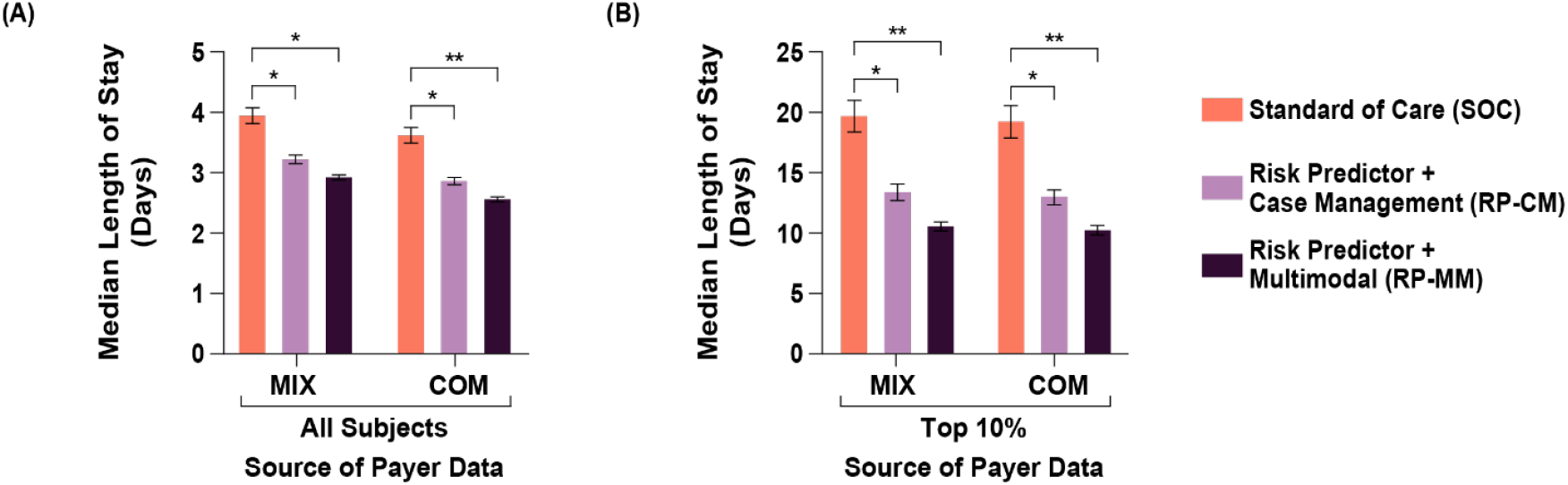
Impact of test-and-treat strategies on neonatal length of hospital stay. The population assessed was either (A) all subjects or (B) those in the top 10% with respect to length of stay. *p < 0.05, determined using Cox proportional hazards regression. **p < 0.001. Abbreviations. COM, U.S. commercial payer dataset; MIX, U.S. state-based dataset with a mix of insurance payers.

**Figure 4.**
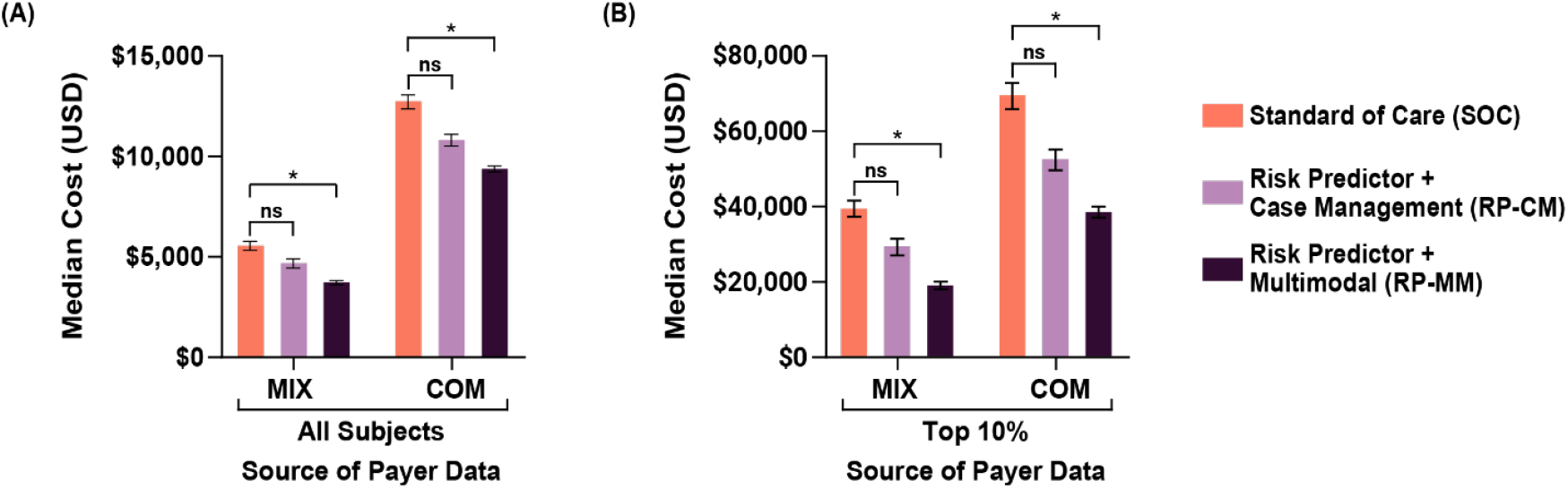
Impact of test-and-treat strategies on neonatal costs. The population assessed was either (A) all subjects or (B) those in the top 10% with respect to costs. \**p* < 0.05, determined using bootstrap intervals. Abbreviations. COM, U.S. commercial payer dataset; MIX, U.S. state-based dataset with a mix of insurance payers; ns, non-significant.

**Figure 5.**
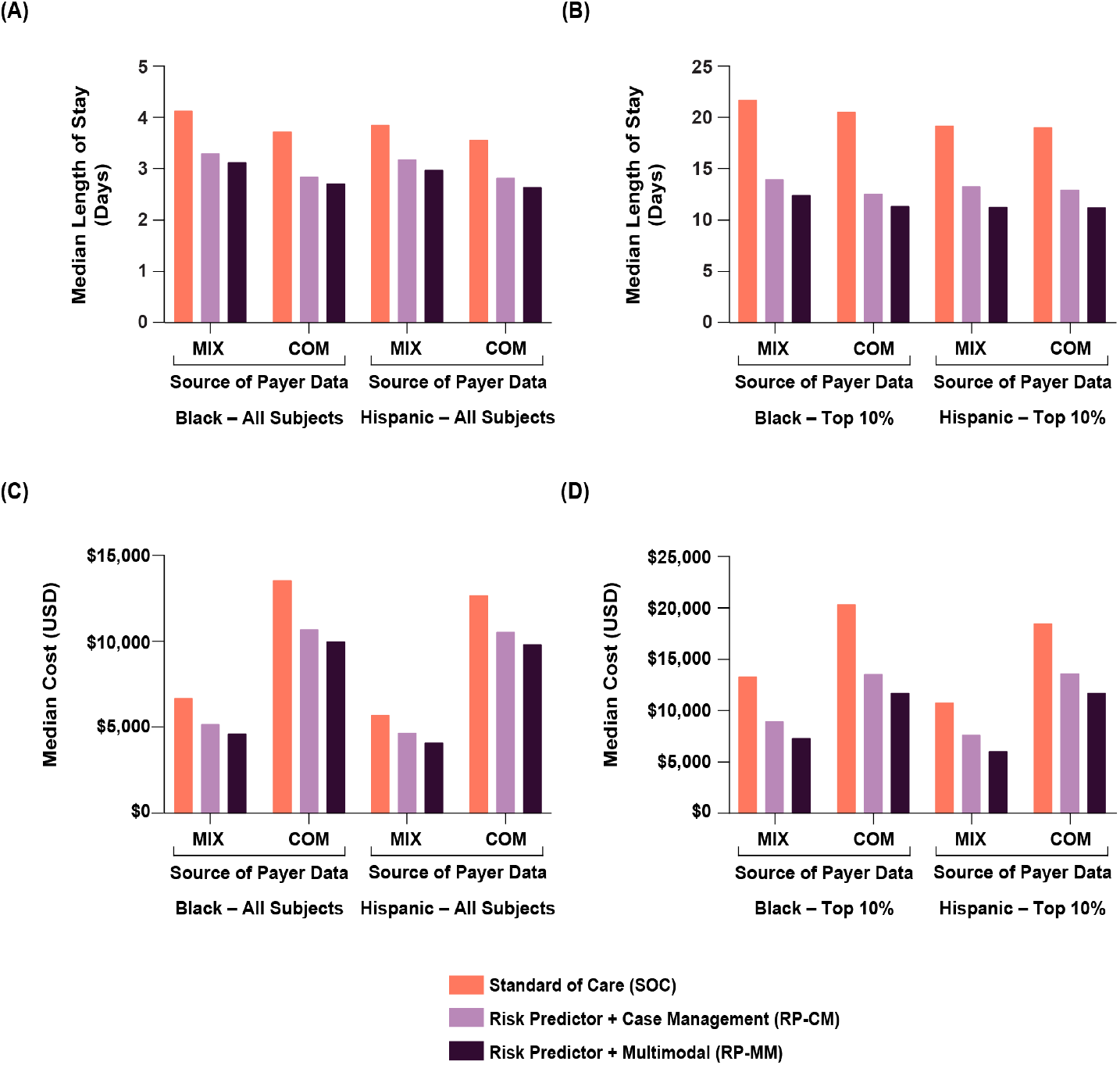
Impact of test-and-treat strategies on neonatal length of hospital stay (A, B) and neonatal costs (C, D) among self-identified Black and Hispanic individuals. The population assessed was either all such subjects (A, C) or those in the top 10% with respect to length of stay or costs (B, D). Abbreviations. COM, U.S. commercial payer dataset; MIX, U.S. state-based dataset with a mix of insurance payers.

Maternal hospital LOS was reduced across all subjects in both the RP-CM (8.5%; IQR, 7-10%) and RP-MM (9.2%, 8-10%) arms using MIX **(Supplemental Table 4)**. For subjects in the top 10% of maternal LOS, stay was reduced by 16% (14-17%) in the RP-CM arm and by 17% (16-19%) in the RP-MM arm. Again, in all cases, the reductions were significant. Maternal LOS point estimates were reduced to a similar degree in Black and Hispanic subgroups as in the study as a whole **(Supplemental Table 5)**.

Neonatal costs’ point estimates decreased non-significantly by 16% (MIX IQR, 9-24%; COM, 10%-20%) in the RP-CM arm. Point estimates of savings after intervention costs were $900 (MIX) and $2,000 (COM) per screened pregnancy. In the RP-MM arm, neonatal costs across all subjects were significantly reduced, by 34% (MIX, 29-38%) and 26% (COM, 23-30%), with median savings after intervention costs of $1,800 (MIX) and $3,300 (COM) per pregnancy screened (**Figure 4** and **Supplemental Table 4**). Among RP-CM subjects in the top 10% of neonatal costs, non-significant point estimate decreases of $10,000 (MIX; 27%; 17-36%) and $17,000 USD (COM; 25%; 16-32%) were seen in cost per screened pregnancy. RP-MM subjects in the top 10% showed significant reductions of $20,000 (MIX, 52%; 47-57%) and $30,000 (COM, 44%; 39-49%) per screened pregnancy. The base and sensitivity cost datasets differed in magnitude of cost reduction, with COM cost reductions being approximately double the MIX cost reductions **(Supplemental Table 4)**. However, conclusions from base and sensitivity cost datasets were retained. Among Black individuals, point estimates of neonatal costs in the base-case scenario (RP-CM) decreased by 21% (MIX, COM) across all subjects and by 31% (MIX) and 34% (COM) in the top 10% (**Figure 5**; **Supplemental Table 5**). Among Hispanic individuals, point estimates of neonatal costs were seen to be reduced by 18% (MIX) and 16% (COM) across all subjects and by 29% (MIX) and 26% (COM) in the top 10%. Again, further improvements were estimated with the addition of pharmacological treatment (RP-MM).

Changes in neonatal morbidity and mortality index scores upon intervention were significant in reduction of the proportion of neonates with scores at or above Level 2 (moderate to severe morbidity or mortality) versus Level 1 (mild morbidity) for both active arms. Reductions in the proportion of neonatal morbidity and mortality Level 2 or higher were 29% (RP-CM) and 41% (RP-MM) (**Table 4**). Comparable decreases also were observed in the Black and Hispanic subgroups, with the proviso that counts of affected babies within subgroups were quite low (**Supplemental Table 6**).

## Discussion

PTB remains the most important complication in obstetrics and neonatology, leading to adverse short- and long-term outcomes for prematurely born neonates. This study assessed the clinical effectiveness and economic benefit of using a risk predictor for identifying higher-risk patients for treatment with well-studied PTB interventions. Our innovative analytical approach combined real-world observational data with simulation of prolonging gestation based on published treatment efficacy to provide real-world effectiveness estimates. We chose several critical metrics to determine whether the modeled studies resulted in improved neonatal outcomes. The results projected significant potential improvements in neonatal morbidity and mortality, neonatal hospital LOS and cost of care. Neonatal improvements occurred without increasing, but rather decreasing, maternal hospital LOS.

The study included a highly diverse racial population, and demonstrated that the benefits of the prematurity prevention strategy seen in the study population as a whole extended to all minority subgroups. While it is clear that the interventions will need to be applied to a larger proportion of the populations in Black and Hispanic women predicted to be at increased sPTB risk, this makes intuitive sense and is clearly worthwhile given the higher known rates of prematurity among these prominent diverse populations. Thus, such a test-and-treat strategy would be promising for groups in this country who are in greatest need of improvement in prematurity-related outcomes.

This study builds upon a previously published modeling study [30], wherein the authors constructed a decision-analytic/Markov cost-effectiveness model evaluating a test-and-treat strategy that used the risk predictor to identify patients at higher risk of PTB, followed by multimodal intervention. These authors found that the test-and-treat strategy was dominant in most scenarios. The current study adds two further assessments: the relationship of risk predictor scores to patient outcomes is based on actual rather than simulated patient data; and a case management intervention is modeled without pharmacological treatments for comparison to multimodal intervention. Overall, the results modeled in the current analysis are consistent with those observed previously.

In efforts toward identifying and treating of at-risk pregnancies with early interventions (progesterone, cervical cerclage, and others) and acute therapies in symptomatic patients (tocolytics, corticosteroids, magnesium sulfate for neuroprotection and antibiotics). Acute maternal administration of antenatal corticosteroids and magnesium sulfate have been demonstrated to improve outcomes associated with premature delivery [35]. Yet the impact of premature delivery has not been materially reduced in the United States. The most consequential cost savings, therefore, are expected when these latter interventions, which can improve clinical outcomes, are applied to the most premature babies.

The current strategy for reducing the consequences of premature delivery is the identification of certain risk factors or specific higher-risk groups and application of approaches that make sense for the etiology associated with their particular risk factor. For small numbers of patients with certain risk factors, an approach such as progesterone therapy for previous preterm delivery or short cervix on endovaginal ultrasound are further examples [4,6,7,36,37]. However, these traditional approaches are limited by the fact that risk factors only capture a small percentage (approximately 11%) [4,5] of patients who will deliver prematurely, whereas the proteomic biomarker captures the majority (75%) of sPTBs [9]. Benefit is therefore more likely, since this approach of risk identification holds promise if it identifies a much larger proportion of those destined to deliver prematurely, and intervention or combination of interventions appropriate for the group chosen are applied [30].

While one cannot necessarily assume efficacy of progesterone in a broader population, a stronger assertion can be made for studies of the potential benefit for case management. Studies in diverse populations have demonstrated reductions in prematurity, shortened NICU stays and/or improvements in neonatal morbidity with programs stratifying higher-risk women to case-management [15-17,38].

Limitations of this study include the impact of reduced adherence to interventions. Further, uncertainty in costs of intervention was not included in the sensitivity analyses. Strengths of this study include the inclusion of real subjects, the prospective application of the risk predictor to blood samples of these subjects and the development of two intervention models independently of subjects of the microsimulation. Although progesterone for use in women with identifiable risk beyond cervical shortening and previous preterm birth is not established and case management has not reached consensus for at risk women, this work suggests that more well-designed controlled studies on these interventions are warranted and have the potential for significant impact. It is important to emphasize that such a risk identification program does not necessarily mean that all those identified to be at higher risk will be amenable to the available interventions to either prevent premature delivery or ameliorate its consequences. For example, those delivering prematurely due to certain medical complications, such as renal failure, or to obstetrical complications not expected to be preventable, such as placental abruption. When and if interventions subsequently become developed that will favorably affect some identifiable cause or causes, then the risk identification approach will become even more valuable, both from improved outcomes and cost-reduction perspectives.

The results of this exercise are important, demonstrating not only the potential benefits for mothers and babies but also the utility of effectiveness assessments combining real-world data and published evidence. Furthermore, this study provides confidence that future studies and real-world use of test-and-treat strategies are worthwhile, ethically justified for pregnant women and are likely to demonstrate positive results where suitable test performance is paired with interventions inclusive of properly implemented stratified case management. As well, this modeling exercise provides a compelling reason to proceed with prospective studies of progesterone and/or case management in patients identified as higher risk with a well-validated risk predictor for PTB. Finally, these results are based on interventions that are currently used in practice, known to be safe and generally found to be acceptable to pregnant women [15,17,35,39].

## Conclusions

High potential exists for the proteomic biomarker risk predictor to be a clinically important component of risk stratification for pregnant women, leading to tangible gains in reducing the impact of PTB.

## Data Availability

Data supporting the results presented here can be requested by emailing the corresponding author. Data will not be made available publicly or in any format that may violate a subject’s right to privacy.

## Transparency

### Author contributions

J.B., G.C.C., J.J.B, and P.E.K conceptualized the study. G.R.M., G.R.S., L.C.L, K.D.H., D.V.C, C.N.S, J.K.B, D.M.H, S.A.L., S.A.S., C.A.M, S.M.W., L.M.P., E.J.S., K.A.B., A.F.H., and A.H.C provided subjects, clinical data, and serum samples for analyses. A.C.F curated data. Formal analyses were done by R.T., J.B. and P.E.K., with methodology designed by J.B, J.J.B, and P.E.K and mentored and reviewed by J.A.F.Z. Code validation was performed by A.D.P. Visualization/data presentation and/or manuscript draft preparation was done by J.B., T.C.F., T.J.G., J.J.B., G.C.C. and P.E.K. The study was supervised by J.J.B. and P.E.K., with P.E.K. serving as project administrator. All authors reviewed, edited, and approved the manuscript.

## Acknowledgments

We wish to acknowledge the study coordinators and research personnel at the study sites, the participants in the TREETOP study and the Sera Prognostics, Inc. clinical laboratory and clinical operations teams. Michael Walker, PhD, reviewed statistical test design and selection. Jennifer Logan, PhD, contributed to the writing of this article.

## Previous presentations

This study was presented as a poster (Abstract #116429) at the ISPOR (The Professional Society for Health Economics and Outcomes Research) annual conference in Washington, DC, on Wednesday May 18, 2022. In addition, a preprint of this manuscript was published on medRxiv and is available at https://doi.org/10.1101/2021.09.08.21262940.

