## Supplemental Data for "Clinical and economic evaluation of a proteomic biomarker preterm birth risk predictor: Cost-effectiveness modeling of prenatal interventions applied to predicted higher-risk pregnancies within a large and diverse cohort"

**SUPPLEMENTAL MATERIAL**
**for Burchard, J. et al.**

**Clinical and economic evaluation of a proteomic biomarker preterm birth risk predictor: Cost-effectiveness modeling of prenatal interventions applied to predicted higher-risk pregnancies within a large and diverse cohort**
https://doi.org/10.1101/2021.09.08.21262940

**Supplemental Table 1.** Probability of neonatal morbidity and mortality level indexed by week of gestation. Proportions of each level of neonatal morbidity and mortality in each week of gestational age at birth were derived from TREETOP observations, with smoothing.

| Gestational age at birth (week) | Level 1 (mild) | Level 2 (moderate) | Level 3 (severe) | Level 4 (death) |
|---|---|---|---|---|
| 23 | 0.0% | 0.0% | 33.0% | 67.0% |
| 24 | 0.0% | 0.0% | 58.9% | 41.1% |
| 25 | 0.0% | 0.0% | 77.4% | 22.6% |
| 26 | 0.0% | 0.0% | 80.8% | 19.2% |
| 27 | 0.0% | 0.0% | 84.0% | 16.0% |
| 28 | 0.0% | 0.0% | 86.0% | 14.0% |
| 29 | 0.0% | 0.0% | 88.0% | 12.0% |
| 30 | 0.0% | 0.0% | 89.4% | 10.6% |
| 31 | 0.0% | 5.9% | 88.2% | 5.9% |
| 32 | 0.0% | 48.4% | 47.6% | 4.0% |
| 33 | 0.0% | 49.7% | 47.1% | 3.3% |
| 34 | 3.9% | 70.6% | 23.5% | 2.0% |
| 35 | 42.1% | 42.1% | 14.0% | 1.8% |
| 36 | 76.4% | 18.0% | 4.5% | 1.1% |
| 37 | 90.9% | 7.9% | 1.0% | 0.3% |
| 38 | 94.7% | 4.4% | 0.7% | 0.2% |
| 39 | 94.5% | 4.7% | 0.7% | 0.1% |
| 40 | 98.1% | 1.7% | 0.3% | 0.0% |
| 41 | 96.1% | 2.5% | 1.3% | 0.1% |
| 42 | 96.1% | 2.5% | 1.3% | 0.1% |

**Supplemental Table 2.** Magnitude of shift in gestational age at birth with modeled interventions for each gestational age week.

| | Weeks | | | | | | | | |
|---|---|---|---|---|---|---|---|---|---|
| **Baseline GA at birth** | 28 | 29 | 30 | 31 | 32 | 33 | 34 | 35 | 36 |
| **RP-CM intervention**<br>**Magnitude of shift in GA at birth** | + 4.1 | + 3.7 | + 3.1 | + 2.5 | + 1.9 | + 1.3 | + 0.9 | + 0.6 | + 0.4 |
| **RP-MM intervention**<br>**Magnitude of shift in GA at birth** | + 7.3 | + 6.5 | + 5.5 | + 4.5 | + 3.5 | + 2.6 | + 1.9 | + 1.3 | + 0.9 |

Abbreviations. GA, gestational age; RP-CM, case management; RP-MM, case management with pharmacological treatment.

**Supplemental Table 3.** Demographic and clinical characteristics of the ACCORDANT study population.

| Subgroup | N (%) | Risk Predictor: Proportion of total showing lower sPTB risk (%) | Risk predictor: Proportion of total showing higher sPTB risk (%) | Higher sPTB risk fold increase over White |
|---|---|---|---|---|
| Black | 173 (20.43) | 19.29 | 22.40 | 1.5 |
| Hispanic | 323 (38.13) | 32.84 | 47.40 | 1.7 |
| Other | 73 (8.62) | 8.62 | 6.17 | 1.0 |
| White | 278 (32.82) | 32.82 | 24.03 | 1.0 |

Abbreviations. sPTB, spontaneous preterm birth.

**Supplemental Table 4.** Impact of test-and-treat strategies on neonatal hospital length of stay (LOS), maternal LOS and neonatal costs. MIX and COM denote the outcome/cost database used for each outcome. The population assessed was either all ACCORDANT subjects (All) or those in the top 10% of each measure (Top 10%). Significant *p*-values are indicated in boldface type.

| Outcome | Active Arm | Population | Median Control | Median Active | % Reduction | Median Difference | *p*-value | IQR Control | IQR Active | IQR % Reduction |
|---|---|---|---|---|---|---|---|---|---|---|
| **MIX - Neonatal LOS** | RP-CM | All | 3.9 | 3.2 | 19% | 0.74 | **0.029**† | 3.8 – 4.1 | 3.1 – 3.3 | 14% – 23% |
| **COM- Neonatal LOS** | RP-CM | All | 3.6 | 2.8 | 22% | 0.79 | **0.0070**† | 3.5 – 3.8 | 2.8 – 2.9 | 17% – 26% |
| **MIX- Maternal LOS** | RP-CM | All | 2.7 | 2.5 | 8.5% | 0.23 | **0.0010**† | 2.7 – 2.8 | 2.5 – 2.5 | 7.2% – 9.7% |
| **MIX - Cost** | RP-CM | All | $5,500 | $4,600 | 16% | $900 | 0.20‡ | $5,300 – $5,700 | $4,300 – $4,800 | 8.9% – 24% |
| **COM - Cost** | RP-CM | All | $13,000 | $11,000 | 16% | $2,000 | 0.098‡ | $12,000 – $13,000 | $10,000 – $11,000 | 10% – 20% |
| **MIX - Neonatal LOS** | RP-CM | Top 10% | 20 | 13 | 33% | 6.5 | **0.024**† | 19 – 21 | 13 – 14 | 25% – 40% |
| **COM - Neonatal LOS** | RP-CM | Top 10% | 19 | 13 | 34% | 6.6 | **0.020**† | 18 – 21 | 12 – 14 | 26% – 40% |
| **MIX - Maternal LOS** | RP-CM | Top 10% | 5.6 | 4.7 | 16% | 0.88 | **0.0041**† | 5.6 – 5.6 | 4.7 – 4.8 | 14% – 17% |
| **MIX - Cost** | RP-CM | Top 10% | $39,000 | $29,000 | 27% | $10,000 | 0.13‡ | $37,000 – $42,000 | $27,000 – $31,000 | 17% – 36% |
| **COM - Cost** | RP-CM | Top 10% | $69,000 | $52,000 | 25% | $17,000 | 0.089‡ | $66,000 – $72,000 | $49,000 – $55,000 | 16% – 32% |
| **MIX - Neonatal LOS** | RP-MM | All | 3.9 | 2.9 | 26% | 1.0 | **0.0018**† | 3.8 – 4.1 | 2.9 – 3.0 | 22% – 29% |
| **COM - Neonatal LOS** | RP-MM | All | 3.6 | 2.5 | 30% | 1.1 | **<0.001**† | 3.5 – 3.7 | 2.5 – 2.6 | 25% – 33% |
| **MIX - Maternal LOS** | RP-MM | All | 2.7 | 2.5 | 9.2% | 0.25 | **<0.001**† | 2.7 – 2.8 | 2.5 – 2.5 | 8.0% – 10% |
| **MIX - Cost** | RP-MM | All | $5,400 | $3,600 | 34% | $1,800 | **0.018** ‡ | $5,200 – $5,700 | $3,500 – $3,700 | 29% – 38% |
| **COM - Cost** | RP-MM | All | $12,000 | $9,200 | 26% | $3,300 | **0.0030**‡ | $12,000 – $13,000 | $9,100 – $9,400 | 23% – 30% |
| **MIX - Neonatal LOS** | RP-MM | Top 10% | 20 | 11 | 46% | 9.0 | **<0.001**† | 18 – 21 | 10 – 11 | 41% – 52% |
| **COM - Neonatal LOS** | RP-MM | Top 10% | 19 | 10 | 47% | 9.0 | **<0.001**† | 18 – 20 | 9.8 – 11 | 41% – 52% |
| **MIX - Maternal LOS** | RP-MM | Top 10% | 5.6 | 4.6 | 17% | 0.97 | **0.0011**† | 5.6 – 5.6 | 4.6 – 4.7 | 16% – 19% |
| **MIX - Cost** | RP-MM | Top 10% | $39,000 | $19,000 | 52% | $20,000 | **0.0040**‡ | $37,000 – $41,000 | $18,000 – $20,000 | 47% – 57% |
| **COM - Cost** | RP-MM | Top 10% | $69,000 | $38,000 | 44% | $30,000 | **0.0010**‡ | $66,000 – $72,000 | $37,000 – $40,000 | 39% – 49% |

†Cox proportional hazards test; significance, $p < 0.05$.
‡Bootstrap test; significance, $p < 0.05$.

Calculations were performed with double-precision floating point numbers. Values were rounded to two significant figures after calculation to reflect the precision of the estimates.

Abbreviations. COM, U.S. commercial payer dataset; IQR, interquartile range; LOS, hospital length of stay; MIX, U.S. state-based dataset with a mix of insurance payers; RP-CM, case management; RP-MM, case management with pharmacological treatment; SoC, standard care control.

**Supplemental Table 5.** Impact of test-and-treat strategies on neonatal hospital length of stay, maternal hospital length of stay and neonatal cost point estimates among self-identified Black and Hispanic individuals. MIX and COM denote the outcome/cost database used for each outcome. The population assessed was either all subjects in the subgroup (All) or those in the top 10% of each measure (Top 10%).

| Outcome | Active Arm | Race/ Ethnicity | Subset | Median Control | Median Active | % Reduction | Median Difference |
|---|---|---|---|---|---|---|---|
| **MIX - Neonatal LOS** | RP-CM | Black | All | 4.1 | 3.3 | 20% | 0.8 |
| **COM- Neonatal LOS** | RP-CM | Black | All | 3.8 | 2.8 | 25% | 0.9 |
| **MIX- Maternal LOS** | RP-CM | Black | All | 2.8 | 2.5 | 8% | 0.2 |
| **MIX - Cost** | RP-CM | Black | All | $6,500 | $5,200 | 21% | $1,400 |
| **COM - Cost** | RP-CM | Black | All | $14,000 | $11,000 | 21% | $2,900 |
| **MIX - Neonatal LOS** | RP-CM | Black | Top 10% | 21 | 14 | 35% | 7.4 |
| **COM- Neonatal LOS** | RP-CM | Black | Top 10% | 21 | 13 | 39% | 8.0 |
| **MIX- Maternal LOS** | RP-CM | Black | Top 10% | 5.1 | 4.7 | 7% | 0.4 |
| **MIX - Cost** | RP-CM | Black | Top 10% | $51,000 | $35,000 | 31% | $16,000 |
| **COM - Cost** | RP-CM | Black | Top 10% | $82,000 | $54,000 | 34% | $28,000 |
| **MIX - Neonatal LOS** | RP-MM | Black | All | 4.1 | 3.1 | 24% | 1.0 |
| **COM - Neonatal LOS** | RP-MM | Black | All | 3.7 | 2.7 | 27% | 1.0 |
| **MIX - Maternal LOS** | RP-MM | Black | All | 2.8 | 2.5 | 9% | 0.2 |
| **MIX - Cost** | RP-MM | Black | All | $6,700 | $4,600 | 31% | $2,100 |
| **COM - Cost** | RP-MM | Black | All | $14,000 | $10,000 | 26% | $3,600 |
| **MIX - Neonatal LOS** | RP-MM | Black | Top 10% | 22 | 12 | 43% | 9.3 |
| **COM - Neonatal LOS** | RP-MM | Black | Top 10% | 20 | 11 | 45% | 9.1 |
| **MIX - Maternal LOS** | RP-MM | Black | Top 10% | 5.1 | 4.7 | 7% | 0.4 |
| **MIX - Cost** | RP-MM | Black | Top 10% | $53,000 | $29,000 | 45% | $24,000 |
| **COM - Cost** | RP-MM | Black | Top 10% | $81,000 | $47,000 | 43% | $34,000 |
| **MIX - Neonatal LOS** | RP-CM | Hispanic | All | 3.8 | 3.2 | 17% | 0.7 |
| **COM - Neonatal LOS** | RP-CM | Hispanic | All | 3.5 | 2.8 | 21% | 0.7 |
| **MIX - Maternal LOS** | RP-CM | Hispanic | All | 2.8 | 2.5 | 10% | 0.3 |
| **MIX - Cost** | RP-CM | Hispanic | All | $5,700 | $4,700 | 18% | $1,000 |
| **COM - Cost** | RP-CM | Hispanic | All | $13,000 | $11,000 | 16% | $2,000 |
| **MIX - Neonatal LOS** | RP-CM | Hispanic | Top 10% | 19 | 13 | 31% | 5.9 |
| **COM - Neonatal LOS** | RP-CM | Hispanic | Top 10% | 19 | 13 | 32% | 6.0 |
| **MIX - Maternal LOS** | RP-CM | Hispanic | Top 10% | 6.3 | 5.0 | 21% | 1.3 |
| **MIX - Cost** | RP-CM | Hispanic | Top 10% | $43,000 | $30,000 | 29% | $12,000 |
| **COM - Cost** | RP-CM | Hispanic | Top 10% | $73,000 | $54,000 | 26% | $19,000 |
| **MIX - Neonatal LOS** | RP-MM | Hispanic | All | 3.8 | 3.0 | 23% | 0.9 |
| **COM - Neonatal LOS** | RP-MM | Hispanic | All | 3.6 | 2.6 | 26% | 0.9 |
| **MIX - Maternal LOS** | RP-MM | Hispanic | All | 2.8 | 2.5 | 11% | 0.3 |
| **MIX - Cost** | RP-MM | Hispanic | All | $5,700 | $4,100 | 28% | $1,600 |
| **COM - Cost** | RP-MM | Hispanic | All | $13,000 | $9,800 | 23% | $2,900 |
| **MIX - Neonatal LOS** | RP-MM | Hispanic | Top 10% | 19 | 11 | 41% | 7.9 |
| **COM - Neonatal LOS** | RP-MM | Hispanic | Top 10% | 19 | 11 | 41% | 7.8 |
| **MIX - Maternal LOS** | RP-MM | Hispanic | Top 10% | 6.3 | 4.9 | 22% | 1.4 |
| **MIX - Cost** | RP-MM | Hispanic | Top 10% | $43,000 | $24,000 | 45% | $19,000 |
| **COM - Cost** | RP-MM | Hispanic | Top 10% | $74,000 | $46,000 | 37% | $27,000 |

Calculations were performed with double-precision floating point numbers. Values were rounded to two significant figures after calculation to reflect the precision of the estimates.

Abbreviations. COM, U.S. commercial payer dataset; IQR, interquartile range; LOS, hospital length of stay; MIX, U.S. state-based dataset with a mix of insurance payers; RP-CM, case management; RP-MM, case management with pharmacological treatment; SoC, standard care control

**Supplemental Table 6.** Impact of test-and-treat strategy on neonatal morbidity and mortality point estimates among self-identified Black and Hispanic individuals. Comparisons were made between the control arm and the two active arms of the proportion of subjects at or below versus above each elevated level of the neonatal morbidity and mortality index. The percent reduction in levels 2 or higher is shown for each active arm.

| Arm | Race/ Ethnicity | Level 1 (mild) | Level 2 (moderate) | Level 3 (severe) | Level 4 (death) | Proportion moderate+ | % Reduction in moderate + |
|---|---|---|---|---|---|---|---|
| **Risk Predictor + Case Management (RP-CM)** | Black | 93.1% | 5.2% | 1.7% | 0.0% | 6.9% | 29.4% |
| **Risk Predictor + Multimodal (RP-MM)** | Black | 93.6% | 5.2% | 1.2% | 0.0% | 6.9% | 35.3% |
| **Standard of Care (SoC)** | Black | 90.2% | 6.9% | 2.3% | 0.6% | 9.8% | – |
| **Risk Predictor + Case Management (RP-CM)** | Hispanic | 93.5% | 3.7% | 2.5% | 0.3% | 6.5% | 22.2% |
| **Risk Predictor + Multimodal (RP-MM)** | Hispanic | 94.4% | 3.4% | 2.2% | 0.0% | 5.6% | 33.3% |
| **Standard of Care (SoC)** | Hispanic | 91.6% | 4.6% | 3.4% | 0.3% | 8.4% | – |

**Supplemental Figure 1.** Functions used to formalize intervention effects defined the means and standard deviations of shifts in gestational (GA) at birth. For each observation $x$, where $x$ is the untreated GA at birth in weeks, calculation using these functions established sigmoid curves relating untreated GA at birth to the magnitude (mean) and variability (standard deviation) of shift in GA at birth with intervention. *max_m* and *max_SD* are the upper asymptotes; *mid_m* and *mid_SD* are the inflection points; and *rate_m* and *rate_SD* are the slopes of the three parameter sigmoid functions.

$$shift\_in\_GA\_at\_birth\_mean(x) = \frac{max\_m}{1 + e^{(mid\_m - x) * rate\_m}}$$

**and**

$$shift\_in\_GA\_at\_birth\_stdev(x) = \frac{max\_SD}{1 + e^{(mid\_SD - x) * rate\_SD}}$$

Abbreviations. GA, gestational age; *m*, mean; *SD*, standard deviation.
